# Newly designed expedited allocation pathways cannot be expected to rely on data that does not currently exist

**DOI:** 10.64898/2026.01.06.26343389

**Authors:** Cameron D. Ekanayake, Syed A. Husain, Miko E. Yu, Joel T. Adler, Chetan Sree Muppavarapu, Jesse D. Schold, Sumit Mohan

## Abstract

Allocation out of sequence (AOOS) allows organ procurement organizations (OPOs) to bypass the standard match-run to expedite kidney placement and prevent nonuse. Inclusion of all AOOS attempts is vital when attempting to assess impact of AOOS on organ utility, including those attempts that do not lead to successful transplant. We assessed the frequency of AOOS documentation in discarded kidneys. Using Scientific Registry of Transplant Recipients (SRTR) Potential Transplant Recipient (PTR) offer data from 2021–2024, we identified match-runs with at least one discarded kidney. AOOS was defined according to Health Resources and Services Administration (HRSA) guidelines and match runs were stratified by kidney recovery and disposition patterns, focusing on 2024 when AOOS was well established. AOOS coding frequency was assessed within each group and across OPOs. In 2024, only 4.3% of all match-runs with at least one discarded kidney contained evidence of AOOS documentation. Across OPOs, AOOS-coded discards ranged from 0.0% to 17.1% (median 3.9%, IQR [2.7–7.6%]). AOOS documentation among discarded kidneys remains rare and inconsistent, suggesting major data-capture deficiencies when attempting to accurately assess AOOS efforts. Improved AOOS reporting is essential before future expedited allocation pathways can be effectively evaluated or implemented.

## 1. Introduction

Kidney transplantation is the preferred treatment for patients with end stage kidney disease, given the dramatic increase in life expectancy and quality of life experienced by recipients. Most patients in the United States who receive a kidney transplant obtain a deceased donor organ that is allocated using a prespecified and objective algorithm that generates a match run, i.e., a list of patients in the order of priority for a given deceased donor organ.^1,2^ Deceased donor organs are offered sequentially to transplant centers in the order that patients are ranked on the match-run. Allocation out of sequence (AOOS) is the circumstance when an organ procurement organization (OPO) deviates from this match run sequence to offer a kidney to a transplant center to expedite placement in an attempt to avoid organ nonuse.^3^ Despite its increasing use, the extent to which AOOS has prevented organ nonuse remains unclear, although there is currently no published evidence of a significant benefit on organ utilization rates.^4-6^ All prior analyses of AOOS have been limited to organs that have been successfully placed with transplant centers and thus does not reflect AOOS efforts for organs that are not utilized.^3-10^ In order to assess the impact of AOOS on organ utilization patterns, it is imperative to consider all cases where AOOS is attempted, regardless of whether the effort to place the organ is successful, especially given that AOOS efforts would likely be greater for organs that were eventually not utilized .^2–4^ We sought to assess the frequency with which AOOS was documented for discarded kidneys to measure the extent of OPOs to allow for successful placement of deceased donor organs with transplant programs.

## 2. Materials and Methods

### 2.1 Data source and cohort construction

This study used data from the Scientific Registry of Transplant Recipients (SRTR). The SRTR data system includes data on all donors, wait-listed candidates, and transplant recipients in the US, submitted by the members of the Organ Procurement and Transplantation Network (OPTN). The Health Resources and Services Administration (HRSA), U.S. Department of Health and Human Services, provides oversight to the activities of the OPTN and SRTR contractors. The data reported here have been supplied by the Hennepin Healthcare Research Institute (HHRI) as the contractor for the Scientific Registry of Transplant Recipients (SRTR). The interpretation and reporting of these data are the responsibility of the author(s) and in no way should be seen as an official policy of or interpretation by the SRTR or the U.S. Government.

We used Potential Transplant Recipient (PTR) kidney offer files for calendar years 2021-2024. Using this file, we identified deceased-donor kidney transplants with a match-run date between January 1, 2021, and December 31, 2024. PTR data was linked to the TX_KI file to confirm organ. If duplicate donor/recipient pairs appeared in multiple match runs, only the first match run was kept, and the rest were excluded. Final organ disposition (transplanted vs discarded) was merged at the organ level and analyses focused only on match runs that were associated with 1 or more discarded kidneys.

### 2.2 Identifying out-of-sequence attempts

Allocation out of sequence (AOOS) was considered present when either (i) a standard AOOS code (861, 862, or 863) was recorded or (ii) an “other, specify” code (799) appeared with AOOS-equivalent language in the accompanying free text (“expedited,” “aggressive,” “open offer,” “out of sequence”).^5^ To assess the relation of AOOS bypasses to acceptance offers within match runs, routine bypass offers were removed but AOOS-related bypasses were retained. We then re-indexed offers within each match run according to their sequential order so that we could identify the position of the first AOOS-coded offer relative to the first accepted offer within the allocation sequence when relevant.

### 2.3 Analysis

Match runs from each year were stratified into three scenarios based on the possible recovery and disposition patterns: Group 1 had match runs with two kidneys recovered and neither was utilized, resulting in bilateral discard. Group 2 included those where only one kidney was recovered but was not transplanted, resulting in a single discard. Group 3 had both kidneys recovered, but only one kidney was transplanted, while the mate kidney was not utilized, resulting in a unilateral discard.

We then identified all AOOS attempts in all match-runs across all 3 groups. In group 3, we were looking for evidence of AOOS for the mate kidney relative to the acceptance of the first kidney. If the first acceptance appeared before an AOOS code in a given match-run, we determined that the accepted kidney was transplanted in sequence, and an AOOS was attempted for the mate kidney. If the AOOS code occurred before the first acceptance, then we assume that the transplanted kidney was AOOS and an attempt to place the mate kidney AOOS was assumed. Given that AOOS reached a peak in 2024 in our dataset with broad awareness, we chose to focus our primary analysis on OPO level coding practices in 2024.

We also examined variations in AOOS efforts to try to avoid an organ discard across OPOs by calculating, for each OPO, the proportion of match runs with discarded kidneys in which any AOOS code was documented. All analyses were performed in Stata 19.0. The Columbia University Irving Medical Center institutional review board approved this study. All research activities were consistent with the principles of the Declaration of Istanbul.

## 3. Results

Between 2021 and 2024, there were 20,544 kidney match runs that had at least 1 discarded kidney. In 2021, 87 (2.0%) of the 4,357 match runs included an AOOS code. In 2022, 171 (3.5%) of the 4,853 match runs had AOOS documentation. The amount of AOOS coding in match runs with discarded kidneys was similar in 2023 and 2024, with AOOS coding occurring in 237 (4.4%) of the 5,410 match runs in 2023 and 254 (4.3%) of the 5,924 match runs in 2024. (Table 1, Figure 1). Kidney-level AOOS coding results for each year are displayed in Table 1.

**Table 1.** Evidence of AOOS placements efforts that were recorded in the PTR file by the OPO for match runs that were associated with kidneys that were discarded. Coding by Donor-Level Stratum, 2021-2024 PTR.

| Donor-Groups | Total Match Runs (n) | 0 AOOS Kidneys | 1 AOOS Kidney | 2 AOOS Kidneys |
| --- | --- | --- | --- | --- |
| **2021** |  |  |  |  |
| Group 1: Two recovered → Bilateral discard | 2,588 | 2,562 (99.0%) | NA | 26 (1.0%) |
| Group 2: Single-kidney recovery → Discard | 235 | 235 (100.0%) | 0 (0.0%) | NA |
| Group 3: Two recovered → Unilateral discard | 1,534 | 1,473 (96.0%) | 7 (0.5%) | 54 (3.5%) |
| **2022** |  |  |  |  |
| Group 1: Two recovered → Bilateral discard | 3,013 | 2,984 (99.0%) | NA | 29 (1.0%) |
| Group 2: Single-kidney recovery → Discard | 209 | 208 (99.5%) | 1 (0.5%) | NA |
| Group 3: Two recovered → Unilateral discard | 1,631 | 1,490 (91.4%) | 20 (1.2%) | 121 (7.4%) |
| **2023** |  |  |  |  |
| Group 1: Two recovered → Bilateral discard | 3,516 | 3,462 (98.5%) | NA | 54 (1.5%) |
| Group 2: Single-kidney recovery → Discard | 225 | 223 (99.1%) | 2 (0.9%) | NA |
| Group 3: Two recovered → Unilateral discard | 1,669 | 1,488 (89.2%) | 27 (1.6%) | 154 (9.2%) |
| **2024** |  |  |  |  |
| Group 1: Two recovered → Bilateral discard | 3,783 | 3,749 (99.1%) | NA | 34 (0.9%) |
| Group 2: Single-kidney recovery → Discard | 257 | 255 (99.2%) | 2 (0.8%) | NA |
| Group 3: Two recovered → Unilateral discard | 1,884 | 1,666 (88.4%) | 20 (1.1%) | 198 (10.5%) |

**Figure 1.**
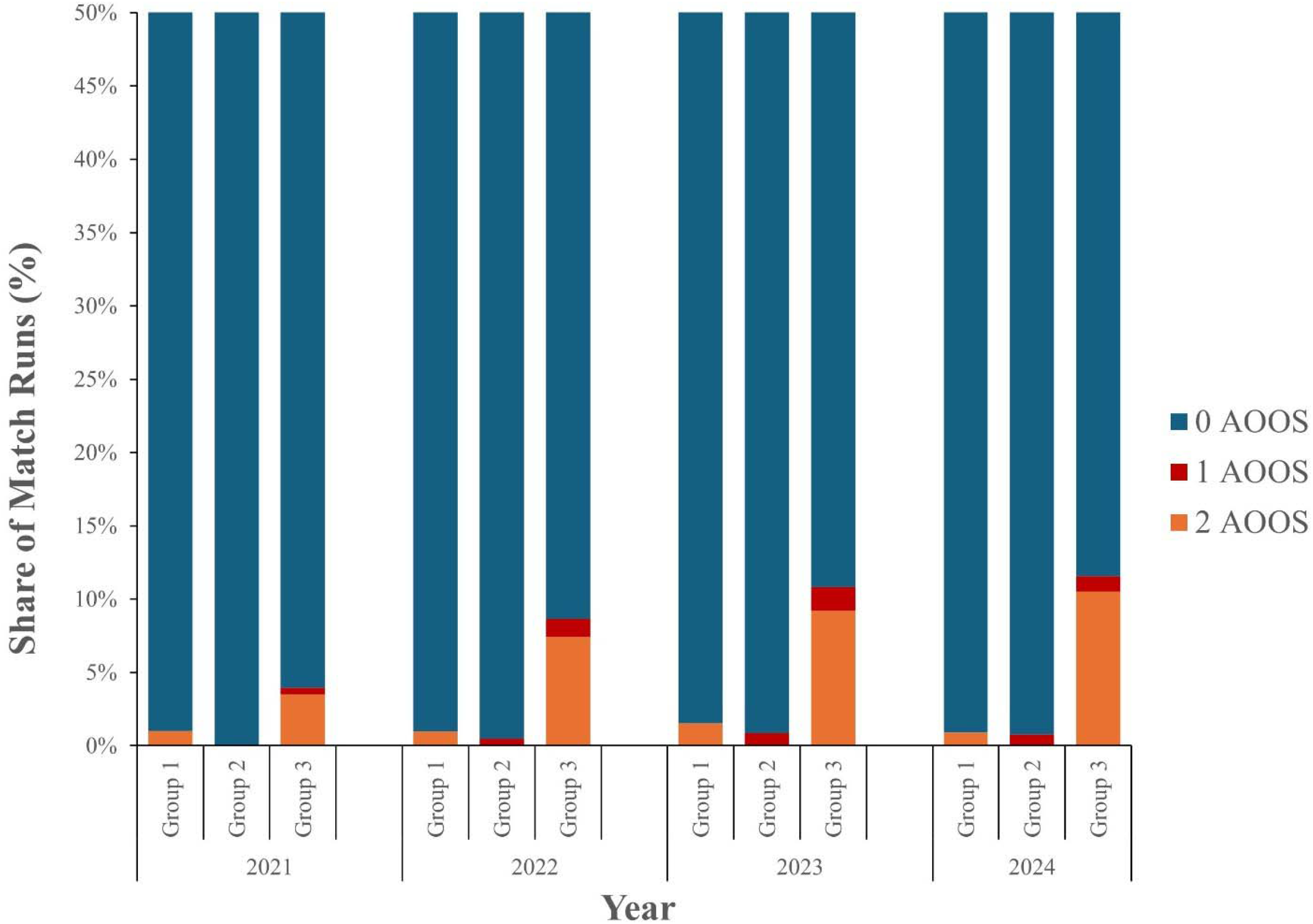
AOOS coding by donor-level stratum, 2021-2024 PTR. ^1^Two recovered → Bilateral discard ^2^Single-kidney recovery → Discard ^3^Two recovered → Unilateral discard

In 2024, fewer than 1% of match runs had an AOOS code included in both groups 1 and 2, i.e., in instances where the match run was not associated with the successful placement of a kidney (Table 1). Among group 3, consisting of match runs in which two kidneys were recovered with one kidney being transplanted and its mate discarded (n = 1,884), 218 of match runs had evidence of an effort to place an organ AOOS. Of these match-runs, 10.5% (n=198) were instances where the first kidney was also placed AOOS while only 1.1% (n =20) of discarded kidneys that followed an in-sequence placement of a kidney had an associated AOOS code – consistent with what was observed in both groups 1 and 2. The absence of AOOS codes in match runs for kidneys that were not successfully placed was similar in the years prior to 2024 (Figure 1). Across OPOs in 2024, the proportion of discarded match runs with evidence of an AOOS attempt ranged from 0.0% to 17.1% (median 3.9%, IQR [2.7–7.6%]). (Figure 2).

**Figure 2.**
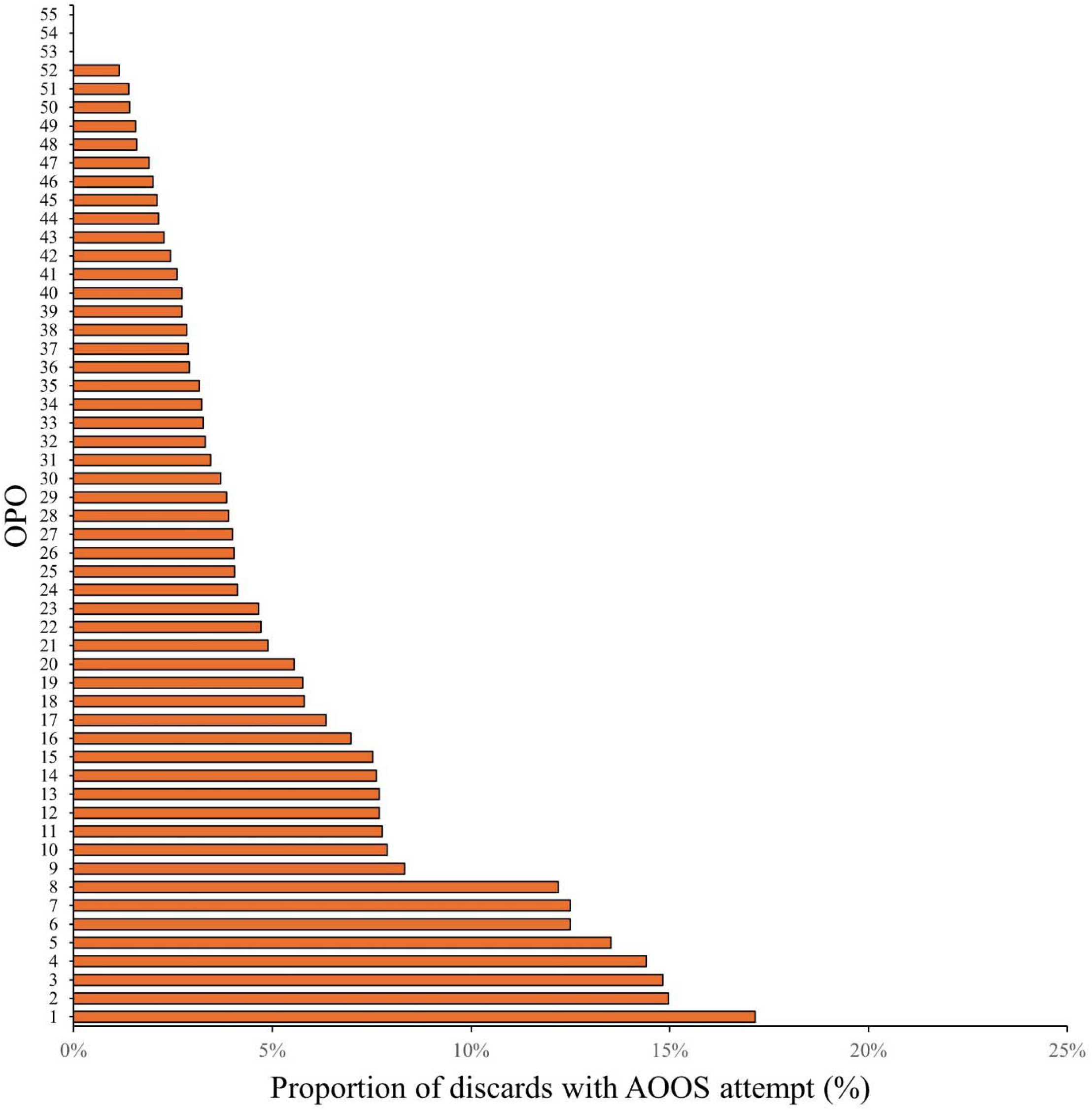
Share of AOOS-coded discarded kidneys by OPO, 2024 PTR.

## 4. Discussion

The absence of out of sequence bypass codes in the overwhelming majority of instances where kidneys are not successfully transplanted raises important questions and considerations for future analyses. Our findings suggest that OPOs’ efforts are not being adequately captured in the data under current reporting guidelines, with over 95% of discarded kidneys in 2024 – the most recent year that data is available - having no evidence of AOOS attempt documented in their respective match-run data. While this represents a small improvement from prior years, it underscores the data challenge that exists.

Furthermore, the frequency of AOOS match runs associated with discarded kidneys varied considerably across OPOs but did not exceed 18% for any OPO, suggesting that this is likely to reflect a data/documentation challenge rather than an absence of effort.^11^

The absence of evidence of AOOS efforts can be the result of one of two possibilities – either there is essentially an absence of accurate data capture of OPO efforts or OPOs are not actually engaging in these efforts for the most difficult to place kidneys that end up being discarded. In the current era of OPO metrics that are reported to be the reason for increasing AOOS efforts, one would expect that it’s more likely that it is the former.

Theoretically, it is possible that an OPO would use AOOS for a less-than-ideal organ when the initial declines are recipient-related while they may chose not to try to place an organ AOOS if there are multiple declines for organ-specific reasons that result in the OPO determining that an AOOS process will also not be successful. While differentiating these would be valuable, the current data precludes us from being able to distinguish these scenarios. As a result, it is difficult, if not impossible, to know when AOOS failed. This hinders the ability to accurately measure the efficacy of AOOS or understand if aggressive transplant centers that receive large numbers of AOOS kidneys are also declining them – and if that organ declination is a result of a surplus supply or true reluctance to use the organ.^12^ Furthermore, the inability to assess process measures such as timing of AOOS initiation, number or the identity of centers contacted with an AOOS offer, or other details precludes the ability to study when AOOS efforts are effective and if there are either organ or operational characteristics that are better associated with a successful placement of the organ. While this would not be the first time that OPTN data capture processes have been found to be lacking, it represents a concerning problem with new policies and allocation pathways being proposed that depend on data elements that are not available.^11,13-16^

Additionally, our study found that there were over 1,800 cases of unilateral discards in just 2024, i.e., where a kidney was discarded while its mate was successfully transplanted, most of which had no AOOS coding. While unilateral discards have been described previously, the reasons for unilateral discards to persist relatively unabated in an era of widespread AOOS raises additional questions about how and when AOOS is being operationalized to lower discards.^17^ For example, a center that is willing to accept one organ from a less-than-ideal donor could have potentially been offered the partner kidney AOOS – or the partner could have been offered to another aggressive center AOOS without accruing additional cold ischemic time. Whether this is happening and centers are reluctant to take both kidneys in that situation cannot be conclusively determined, given the dataset in its current form. It is important to note that the entry of refusal codes or AOOS codes is not necessarily a real time process with codes being entered often days later. Time stamps appear to be related to the time of the data entry as opposed to the time of the actual offer notification or its acceptance/decline which adds additional layers of complexity when attempting to understand the temporal sequence of events.^18,19^

Finally, we note that without being able to adequately identify failed AOOS attempts, our ability to measure how effective AOOS is at preventing discards is further limited – especially given that at the present time there is no evidence available to suggest that AOOS is effective at lowering discard rates.^5^ While there is increased interest in the creation of expedited pathways for less-than-ideal organs, the absence of these data would suggest the need for clear guidelines that are transparent and associated with robust data capture so that we can iteratively improve deceased donor allocation in the United States. Newly designed expedited allocation pathways cannot be expected to rely on data that does not exist currently.

## Data Availability Statement

The data used for this analysis are available upon request to the SRTR.

## Acknowledgements

The data reported here have been supplied by the Hennepin Healthcare Research Institute (HHRI) as the contractor for the Scientific Registry of Transplant Recipients (SRTR). The interpretation and reporting of these data are the responsibility of the author(s) and in no way should be seen as an official policy of or interpretation by the SRTR or the U.S. Government.

## Abbreviations

AOOS: 
HHRI: 
OPO: 
OPTN: 
PTR: 
SRTR: 

## References

1. Puttarajappa CM, Hariharan S, Zhang X, et al. Early Effect of the Circular Model of Kidney Allocation in the United States. J Am Soc Nephrol. Jan 1 2023;34(1):26–39. doi:10.1681/asn.2022040471

2. Mohan S, Yu M, Husain SA. Equity and the operational considerations of the kidney transplant allocation system. Curr Opin Organ Transplant. Apr 1 2025;30(2):146–151. doi:10.1097/MOT.0000000000001201

3. King KL, Husain SA, Perotte A, Adler JT, Schold JD, Mohan S. Deceased donor kidneys allocated out of sequence by organ procurement organizations. Am J Transplant. May 2022;22(5):1372– 1381. doi:10.1111/ajt.16951

4. Adler JT, Cron DC, Kuk AE, et al. Association between out-of-sequence allocation and deceased donor kidney nonuse across organ procurement organizations. Am J Transplant. Feb 17 2025;doi:10.1016/j.ajt.2025.02.005

5. Mohan S, Yu ME, Adler JT, et al. Out-of-Sequence Placement of Deceased Donor Kidneys is Exacerbating Inequities in the United States. Clin J Am Soc Nephrol. Sep 24 2025;doi:10.2215/CJN.0000000869

6. Cron DC, Husain SA, Kuk AE, Mohan S, Adler JT. Increasing Incidence of Out-of-Sequence Allocation of Deceased-Donor Kidneys. Kidney 360. 2024:10.34067/KID.0000000640. doi:10.34067/kid.0000000640

7. Yu M, Husain SA, Adler JT, Maclay L, Schold JD, Mohan S. Out-of-Sequence Placement of Deceased Donor Kidneys Is Exacerbating Inequities. J Am Soc Nephrol. Oct 2024 2024;35(10s):B11.

8. Mohan S, Yu ME, Adler JT, et al. Socioeconomic Disparities in Out-of-Sequence Placement of Deceased Donor Kidneys in the US. JAMA Intern Med. Jul 7 2025;doi:10.1001/jamainternmed.2025.2783

9. Tucker EG, Yu ME, Adler JT, et al. Underrecognition of deceased donor kidney out-of-sequence allocation due to increasing use of free-text coding. Am J Transplant. Apr 9 2025;doi:10.1016/j.ajt.2025.04.002

10. Liyanage LN, Akizhanov D, Patel SS, et al. Contemporary prevalence and practice patterns of out-of-sequence kidney allocation. Am J Transplant. Feb 2025;25(2):343–354. doi:10.1016/j.ajt.2024.08.016

11. Tsapepas DS, King K, Husain SA, et al. UNOS Decisions Impact Data Integrity of the OPTN Data Registry. Transplantation. Dec 1 2023;107(12):e348–e354. doi:10.1097/TP.0000000000004792

12. Li MT, King KL, Husain SA, Schold JD, Mohan S. Deceased Donor Kidneys Utilization and Discard Rates During COVID-19 Pandemic in the United States. Kidney Int Rep. Sep 2021;6(9):2463– 2467. doi:10.1016/j.ekir.2021.06.002

13. Tsapepas D, King KL, Husain SA, Mohan S. Evaluation of kidney allocation critical data validity in the OPTN registry using dialysis dates. Am J Transplant. Jan 2020;20(1):318–319. doi:10.1111/ajt.15616

14. Yu M, King KL, Husain SA, et al. Discrepant Outcomes between National Kidney Transplant Data Registries in the United States. J Am Soc Nephrol. Aug 3 2023;doi:10.1681/ASN.0000000000000194

15. Noreen SM, Patzer RE, Mohan S, et al. Augmenting the Unites States transplant registry with external mortality data: A moving target ripe for further improvement. Am J Transplant. Feb 2024;24(2):190–212. doi:10.1016/j.ajt.2023.09.002

16. Yu K, King K, Husain SA, Mohan S. Variations in Deceased Donor Terminal Creatinine Values Reported in the OPTN Data Registry. Clin J Am Soc Nephrol. Apr 2022;17(4):565–567. doi:10.2215/CJN.15511121

17. Husain SA, Chiles MC, Lee S, et al. Characteristics and performance of unilateral kidney transplants from deceased donors. Clin J Am Soc Nephrol. 2018;13(1):118.

18. Kilambi V, Barah M, Formica RN, Friedewald JJ, Mehrotra S. Evaluation of Opening Offers Early for Deceased Donor Kidneys at Risk of Nonutilization. Clin J Am Soc Nephrol. 2023:10.2215.

19. Cron DC, Husain SA, King KL, Mohan S, Adler JT. Increased volume of organ offers and decreased efficiency of kidney placement under circle-based kidney allocation. Am J Transplant. Aug 2023;23(8):1209–1220. doi:10.1016/j.ajt.2023.05.005

